# A Real-World Pharmacovigilance Study of FDA Adverse Event Reporting System (FAERS) for Fedratinib

**DOI:** 10.64898/2025.12.30.25343201

**Authors:** Lingjun Wu, Yazhou Wang, Duanfeng Jiang, Lihua Tian, Ping Zou, Jingjie Liao, Qiaoyun Zhang, Yanyuan Li, Yuke Xie, Yushen Huang

## Abstract

**Background:** Using the FDA Adverse Event Reporting System (FAERS) database, this pharmacovigilance investigation systematically assessed the adverse events (AEs) associated with Fedratinib use in real-world clinical practice.

**Methods:** Using the FAERS database, we performed a disproportionality analysis incorporating four distinct signal detection methodologies: ROR, PRR, BCPNN, and MGPS. Subgroup analyses were conducted to evaluate the effects of age and gender on Fedratinib-associated AEs. Furthermore, a time-to-onset analysis was performed to characterize the temporal patterns of AEs.

**Results:** The FAERS database comprised 10,011,422 individual case safety reports, of which 1,284 were classified as Fedratinib-related AEs, encompassing 38 significant preferred terms (PTs). The most frequently reported adverse reactions included diarrhoea, nausea, vomiting, constipation, abdominal discomfort, fatigue, anaemia, platelet count decreased, and Wernicke’s encephalopathy. New AEs emerged from the study, such as blood potassium increased, hyperkalaemia, gout, hypocalcaemia, renal failure, renal disorder, and gallbladder disorder. Higher rates were observed in males over 65 years of age. Most cases occurred within the first month of Fedratinib treatment, with the incidence of related AEs decreasing over time.

**Conclusion:** This current study marks the debut investigation regarding Fedratinib’s safety in actual clinical practice, which providing substantive evidence to inform future pharmacovigilance investigations of this drug.

## 1. Introduction

Myelofibrosis, a BCR-ABL1-negative myeloproliferative neoplasm (MPN), is pathologically characterized by clonal bone marrow proliferation, progressive fibrosis, cytopenias, extramedullary hematopoiesis with consequent hepatosplenomegaly, elevated proinflammatory cytokines, and constitutional symptoms[1]. Currently, clinical treatments for MF include JAK kinase inhibitors, immunomodulators, and erythropoietin[2]. JAK inhibitors remain the main treatment option for patients with MF, which meet the clinical needs of this disease for different indications[3,4]. At the same time, clinicians are facing increasingly complex problems in the selection and proper use of these drugs.

Fedratinib, a selective inhibitor of JAK2 and FLT3 kinases, was approved by the FDA on August 16, 2019, for the treatment of intermediate-2 or high-risk MF, including both primary MF and secondary MF (arising from polycythemia vera or essential thrombocythemia) in adult patients. Clinical trial data support the use of Fedratinib as a second-line therapy in patients with cytopenia accompanied by either clinically significant splenomegaly or debilitating systemic manifestations. However, particular clinical vigilance is required for Fedratinib use in malnourished patients or those with gastrointestinal symptoms, necessitating intensive thiamine monitoring as a preventive measure against Wernicke’s encephalopathy (WE) development[3]. Although the drug was previously required by the FDA to carry a ‘black box warning’ [5] and clinical trials were temporarily suspended due to potential risks of severe encephalopathy (including WE), subsequent studies confirmed no direct causal relationship between WE and Fedratinib[6]. The above results suggest that it is particularly important to evaluate this drug in the real world after its market launch.

Phase II/III clinical trials and randomized controlled studies have consistently demonstrated that the most frequent adverse drug reactions associated with Fedratinib administration include anaemia, thrombocytopenia, gastrointestinal symptoms (diarrhoea, nausea, and vomiting), and abnormal liver function. Among them, anaemia and thrombocytopenia are grade 3-4 adverse reactions, and most of the other adverse reactions are grade 1-2[7–10]. Novel JAK inhibitors and combination approaches are being explored to overcome these limitations. Consequently, given these findings, the systematic employment of pharmacovigilance data mining approaches is essential to uncover potential Fedratinib-related adverse drug reactions in routine clinical environments. However, most studies on Fedratinib to date have centered around preclinical study. Considering the extensive clinical utilization of Fedratinib alongside the limited real-world evidence regarding its AEs, this study systematically analyzed real-world pharmacovigilance data to characterize the safety profile.

The FAERS represents one of the most extensive global pharmacovigilance databases currently available and can be used to monitor AEs related to drug use[11]. The database is an open voluntary reporting system for real adverse event reports filed by medical practitioners, pharmacists, manufacturers, and other individuals[12]. Given the paucity of real-world evidence regarding Fedratinib-associated AEs, this study systematically evaluated Fedratinib-associated AEs through an analysis of FAERS data from 2019 onward, providing valuable evidence to inform its rational clinical use.

## 2. Materials and Methods

### 2.1 Data source and processing

FAERS provides the FDA with an essential pharmacovigilance tool for the ongoing safety evaluation of approved drugs and therapeutic biologics through adverse event monitoring. The FAERS database is structurally organized into seven discrete datasets that collectively facilitate comprehensive pharmacovigilance analyses: (1) demographic and administrative information (DEMO), documenting patient characteristics and case management details; (2) adverse drug reaction reports (REAC), capturing all documented AEs; (3) patient outcomes (OUTC), recording clinical consequences of reported events; (4) drug exposure data (DRUG), detailing all medications involved; (5) therapy duration records (THER), specifying treatment initiation and cessation dates; (6) reporter information (RPSR), identifying submission sources; and (7) indication/diagnosis documentation (INDI), providing clinical context for medication use. Given the quarterly update cycle of the FAERS database, duplicate reports were identified and removed using the standardized FDA deduplication algorithms. For enhanced analytical accuracy and reliability, we extracted the PRIMARYID, CASEID, and FDA_DT fields from the DEMO table. The records were then sorted chronologically by CASEID, FDA_DT, and PRIMARYID, retaining only the most recent FDA_DT entry for each unique CASEID to ensure data uniqueness and to prevent duplication. When duplicate records shared identical CASEID and FDA_DT values, the entry with the higher PRIMARYID was prioritized for the analysis. All reports flagged for removal in the FDA’s quarterly deletion updates were systematically excluded from our dataset[13]. Medications reported in the FAERS database were categorized according to standard pharmacovigilance classifications: primary suspect (PS), secondary suspect (SS), concomitant (C), and interacting (I) drugs. For enhanced data accuracy, we exclusively included cases with Fedratinib coded as the PS medication. Finally, a systematic analysis of the FAERS database identified 10,011,422 case reports involving Fedratinib exposure. After excluding duplicates, 8,528,344 reports showed Fedratinib as a PS drug, resulting in 1,284 Fedratinib-associated AEs and 2,670 Fedratinib-associated PTs. Figure 1 presents the complete workflow of our systematic screening methodology.

**Figure 1.**
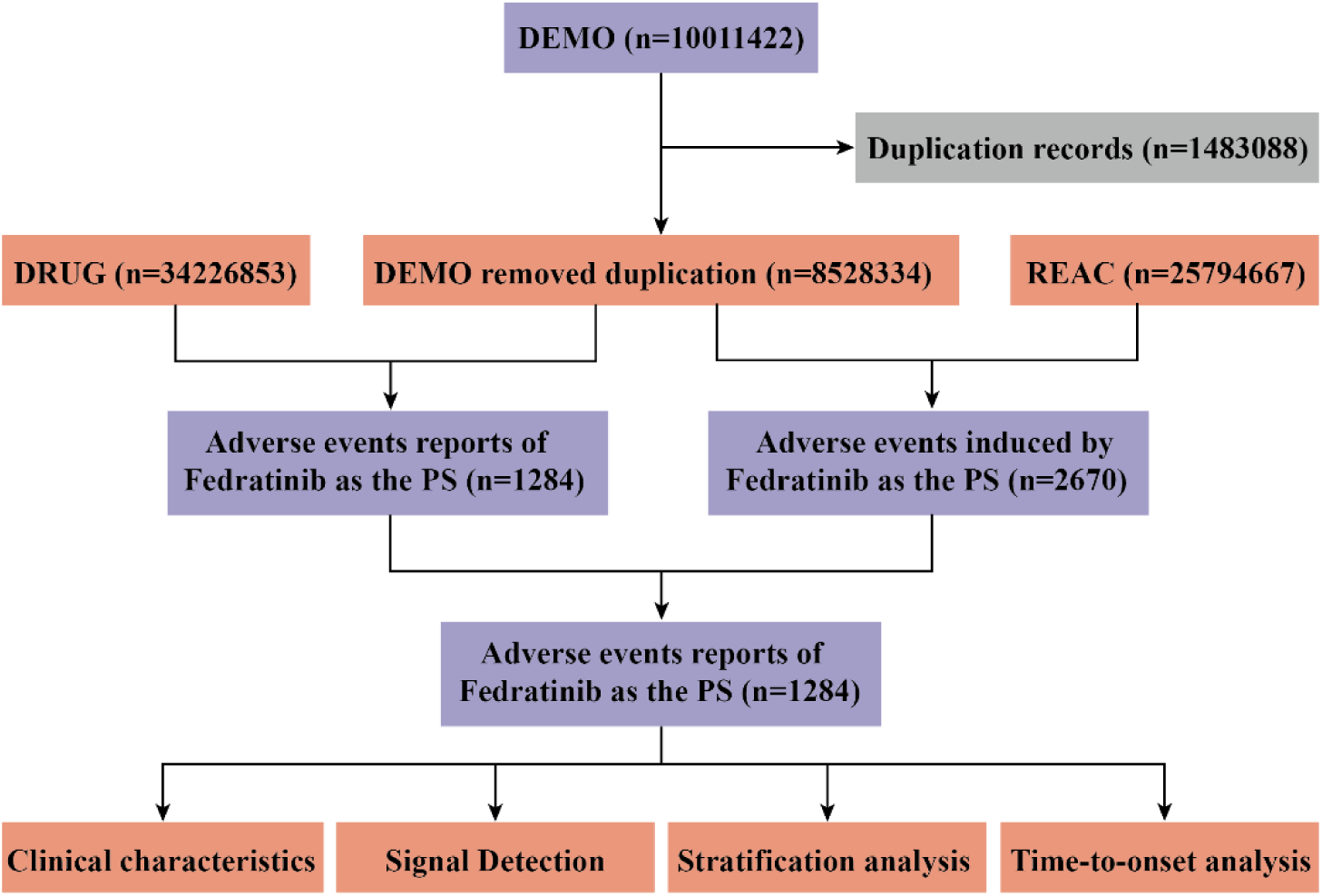
Flow diagram of this study (DEMO: demographic and administrative information, DRUG: drug information, REAC: preferred terminology for adverse events, PS: primary suspect drug).

### 2.2 Data analysis

Disproportionality analysis represents a widely utilized methodology in pharmacovigilance investigations, by assuming that there may be a causal relationship between the drug and the adverse reaction, and conducting subsequent clinical evaluations on potential case reports. Data mining based on quadruple tables, we applied four algorithms, reporting odds ratio (ROR) [14], proportional reporting ratio (PRR) [15], bayesian confidence propagation neural network (BCPNN)[16], and multi-item gamma poisson shrinker (MGPS)[17], to perform disproportionate analysis to identify signals of potentially increased risk of Fedratinib-associated AEs. ROR and PRR represent frequentist (non-Bayesian) methodologies that exhibit high sensitivity but limited specificity. In contrast, the BCPNN and MGPS approaches are more appropriate for analyzing complex variables, albeit with reduced sensitivity. Under conditions of limited or incomplete data availability, BCPNN can still perform effective signal detection, and the MGPS algorithm demonstrates superior performance in detecting rare adverse reaction signals. A positive correlation was observed between the magnitudes of these four parameters and the signal strength, with higher values indicating more robust detection signals. The integrated application of four distinct algorithms in this study capitalizes on their complementary advantages, thereby broadening the detection scope, enabling multi-perspective validation, and ultimately enhancing the stability, comprehensiveness, and reliability of the research outcomes[14,18]. For systematic characterization and analysis of adverse drug events, all cases in our database were coded according to PTs and subsequently mapped to the highest-level SOC within the Medical Dictionary for Regulatory Activities (MedDRA) hierarchy (version 26.0)[18]. The AEs signals selected for analysis in our study satisfied all four algorithmic criteria. The detailed formulations and threshold values of the four algorithms are summarized in Table 1.

**Table 1.**
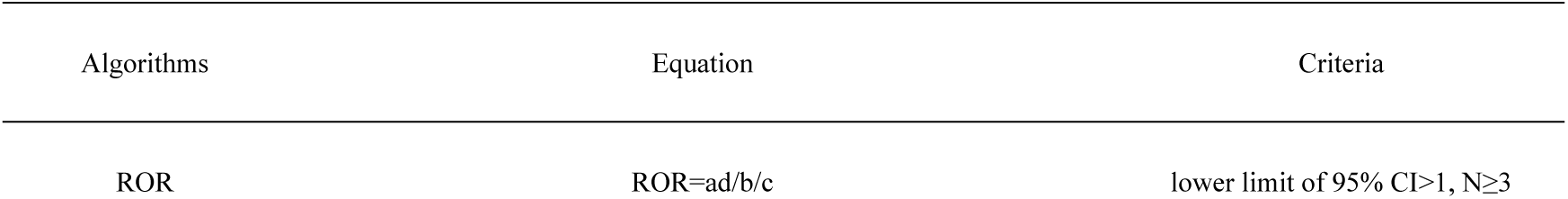
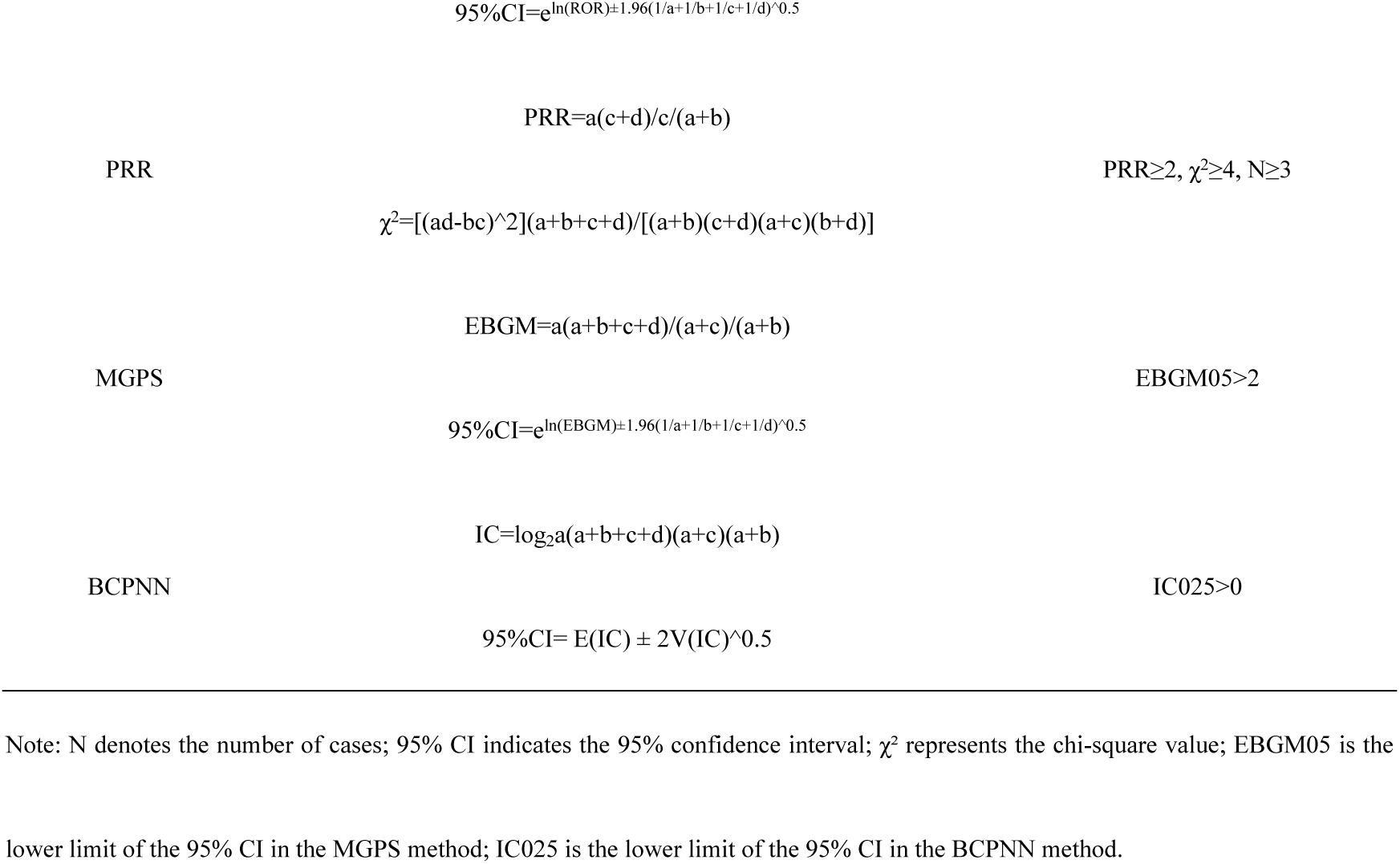
Formulas and criteria for disproportionality analysis.

AEs meeting any of the following criteria: life-threatening or resulting in hospitalization, disability, or death, were categorized as SAEs. Reported AEs were stratified by severity into SAE and NSAE groups. Subsequent subgroup analyses examined potential demographic variations, including age stratification (≤65 vs. >65 years) and gender distribution (male/female), to evaluate the demographic influences on adverse event severity. To ensure methodological accuracy, we systematically excluded all reports with incomplete age or gender records.

The time-to-onset was computed as the duration between EVENT_DT (adverse event onset date) and START_DT (Fedratinib therapy initiation date). The data exclusion criteria comprised records with chronologically inconsistent or incomplete date entries and cases where EVENT_DT preceded START_DT. The incidence of AEs depends on the pharmacological mechanism of the drug and demonstrates temporal variability[19]. The Weibull shape parameter (WSP) analysis quantifies temporal variations in adverse event incidence, delineating whether the associated risk demonstrates an increasing, decreasing, or constant trend over the treatment duration[20]. Two fundamental parameters govern the Weibull distribution: α (scale parameter) and β (shape parameter), which collectively determine its statistical properties. In this investigation, we focused exclusively on parameter β for analysis and interpretation. Three distinct temporal risk patterns were identified based on β values and their 95% confidence intervals (CIs): (1) early failure pattern (β < 1 with 95% CI < 1), indicating a decreasing risk of AEs over time; (2) random failure pattern (β ≈ 1 with 95% CI encompassing 1), suggesting a constant risk throughout the observation period; and (3) wear failure pattern (β > 1 with 95% CI excluding 1), demonstrating an increasing risk trend with prolonged exposure[21,22].

## 3. Results

### 3.1 Descriptive characteristics

Table 2 summarizes the clinical characteristics of the Fedratinib-associated AEs. The FAERS database contained 1,284 reportable events that met our inclusion criteria. Males (51.17%) reported a higher proportion of AEs than females (45.56%), and the proportion of unknown gender was 3.27%. Age distribution analysis revealed a predominance of AEs in elderly patients (>65 years, 49.07%), with significantly higher incidence rates than in younger adults (18-65 years, 15.18%). Of the reported outcomes, the most common was hospitalization (233, 18.15%), followed by other serious outcomes (213,16.59%), death (176,13.71%), life-threatening (15,1.17%), and disability (5,0.39%). Geographically, the highest reporting frequencies were observed in five nations: the United States, Canada, France, Italy, and the United Kingdom, with the United States having the highest percentage of reports (1,043,81.23%). Most reports are submitted by consumers and health professionals, with approximately the same number of reports submitted by medical doctors and pharmacists. The yearly distribution shows that the highest number of reports was recorded in 2021 (322,25.08%), with a rapid increase in the number of reports from 2019 (3.19%) to 2021 (25.08%), and a decreasing trend from 2021 onwards to 2025. The percentage of both serious and non-serious reports of ADEs caused by Fedratinib was 50.00%, which shows that all AEs accompanying the use of Fedratinib deserve extra attention.

**Table 2.**
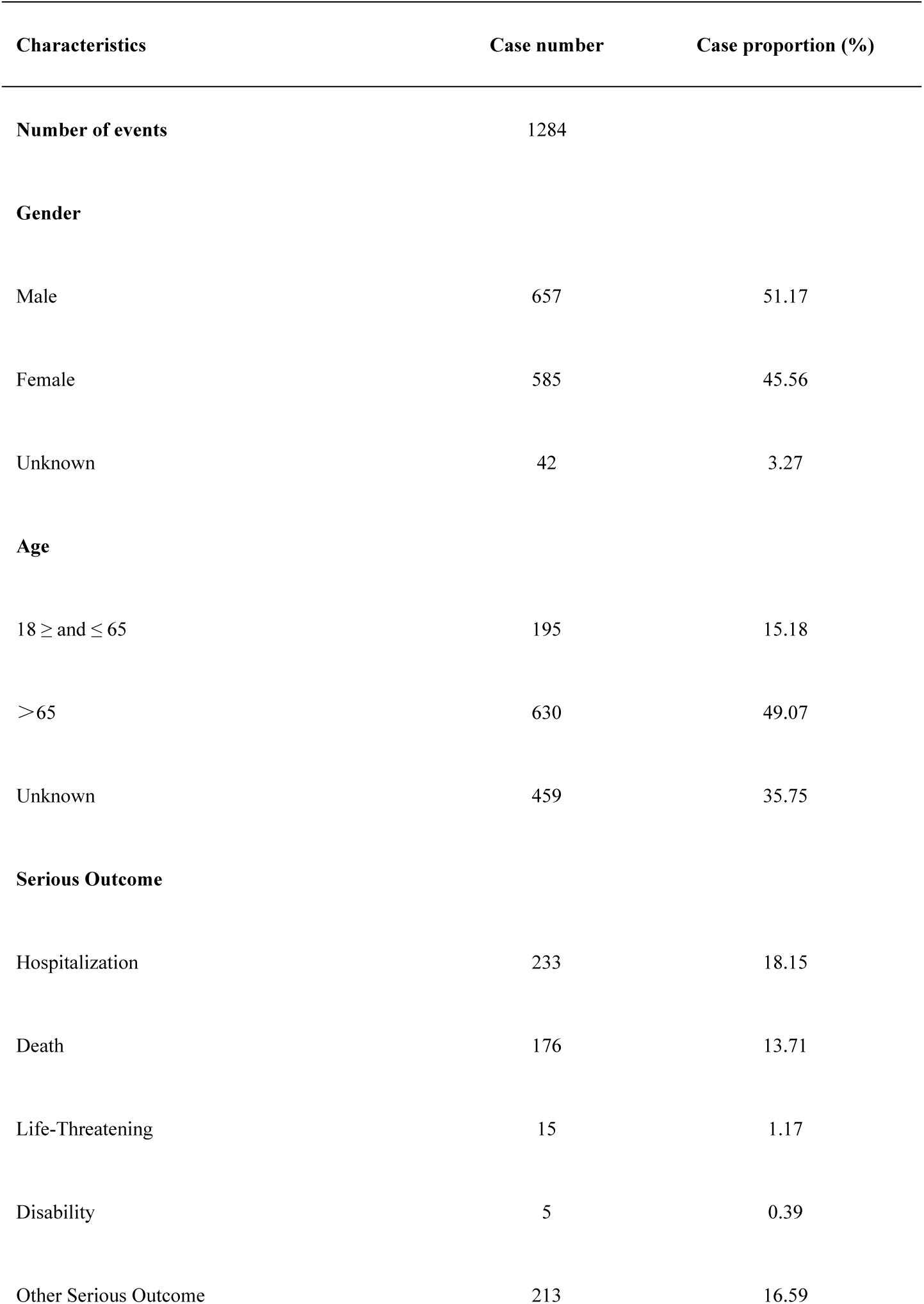
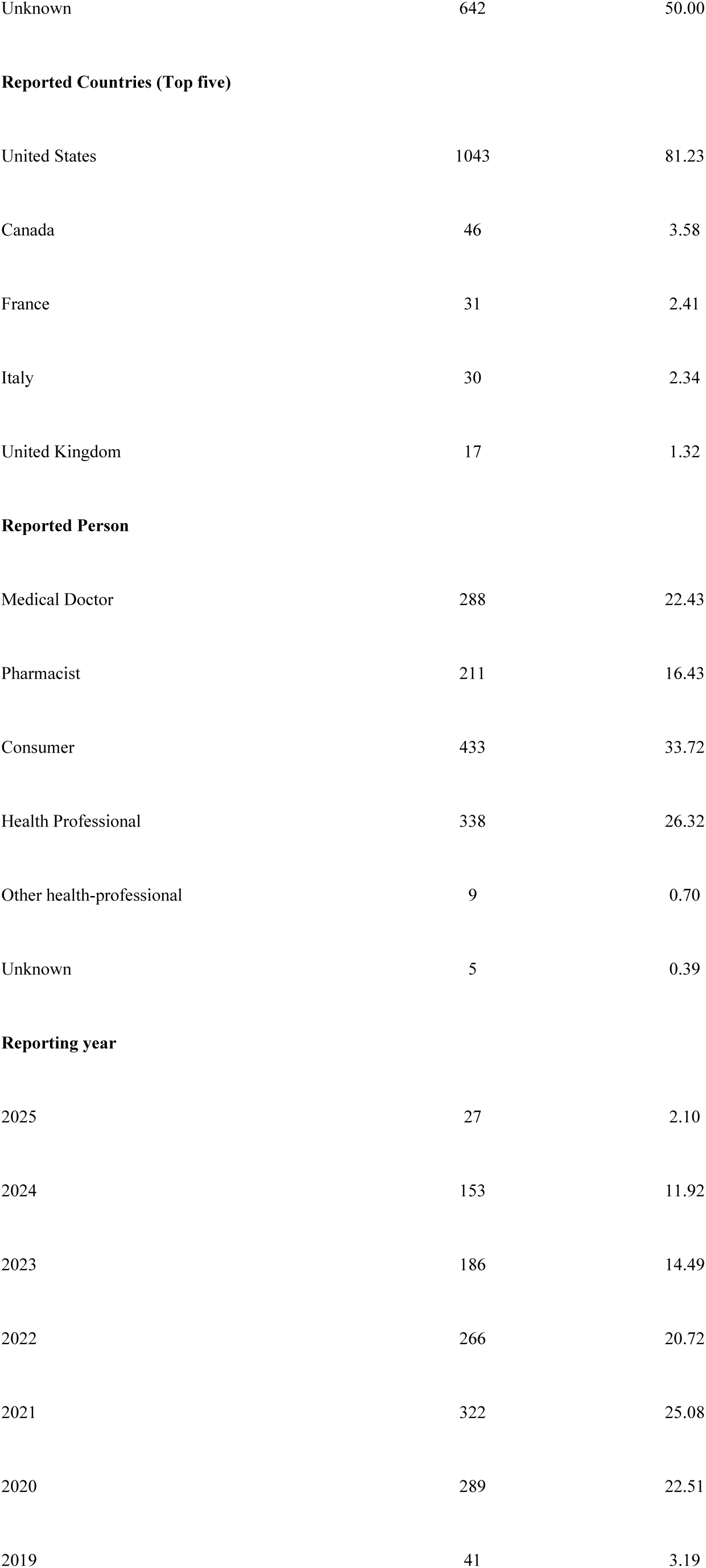
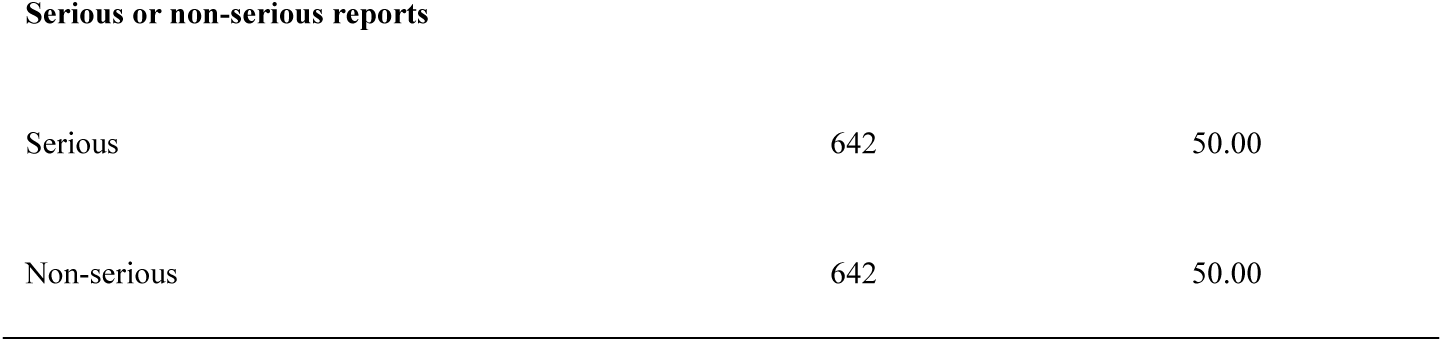
Clinical characteristics of reports with Fedratinib from the FAERS database (2019 to 2025)

| Characteristics | Case number | Case proportion (%) |
| --- | --- | --- |
| **Number of events** | 1284 |  |
| **Gender** |  |  |
| Male | 657 | 51.17 |
| Female | 585 | 45.56 |
| Unknown | 42 | 3.27 |
| **Age** |  |  |
| 18 ≥ and ≤ 65 | 195 | 15.18 |
| >65 | 630 | 49.07 |
| Unknown | 459 | 35.75 |
| **Serious Outcome** |  |  |
| Hospitalization | 233 | 18.15 |
| Death | 176 | 13.71 |
| Life-Threatening | 15 | 1.17 |
| Disability | 5 | 0.39 |
| Other Serious Outcome | 213 | 16.59 |
| Unknown | 642 | 50.00 |
| **Reported Countries (Top five)** |  |  |
| United States | 1043 | 81.23 |
| Canada | 46 | 3.58 |
| France | 31 | 2.41 |
| Italy | 30 | 2.34 |
| United Kingdom | 17 | 1.32 |
| **Reported Person** |  |  |
| Medical Doctor | 288 | 22.43 |
| Pharmacist | 211 | 16.43 |
| Consumer | 433 | 33.72 |
| Health Professional | 338 | 26.32 |
| Other health-professional | 9 | 0.70 |
| Unknown | 5 | 0.39 |
| **Reporting year** |  |  |
| 2025 | 27 | 2.10 |
| 2024 | 153 | 11.92 |
| 2023 | 186 | 14.49 |
| 2022 | 266 | 20.72 |
| 2021 | 322 | 25.08 |
| 2020 | 289 | 22.51 |
| 2019 | 41 | 3.19 |
| Serious or non-serious reports |  |  |
| Serious | 642 | 50.00 |
| Non-serious | 642 | 50.00 |

### 3.2 Signal detection

According to the statistics, Figure 2 presents the distribution of Fedratinib-related case reports across SOCs. 25 SOCs were involved in Fedratinib-induced AEs. The five most frequently affected SOCs were general disorders and administration site conditions (n=552,20.67%), gastrointestinal disorders (n=482,18.05%), investigations (n=317,11.87%), nervous system disorders (n=164,6.14%), injury, poisoning and procedural complications (n=132,11.87%). We found that the PTs under 13 of the 25 SOCs met the criteria, which were gastrointestinal disorders, general disorders and administration site conditions, investigations, blood and lymphatic system disorders, metabolism and nutrition disorders, renal and urinary disorders, cardiac disorders, musculoskeletal and connective tissue disorders, nervous system disorders, skin and subcutaneous tissue disorders, eye disorders, psychiatric disorders, and hepatobiliary disorders (Table 3).

**Figure 2.**
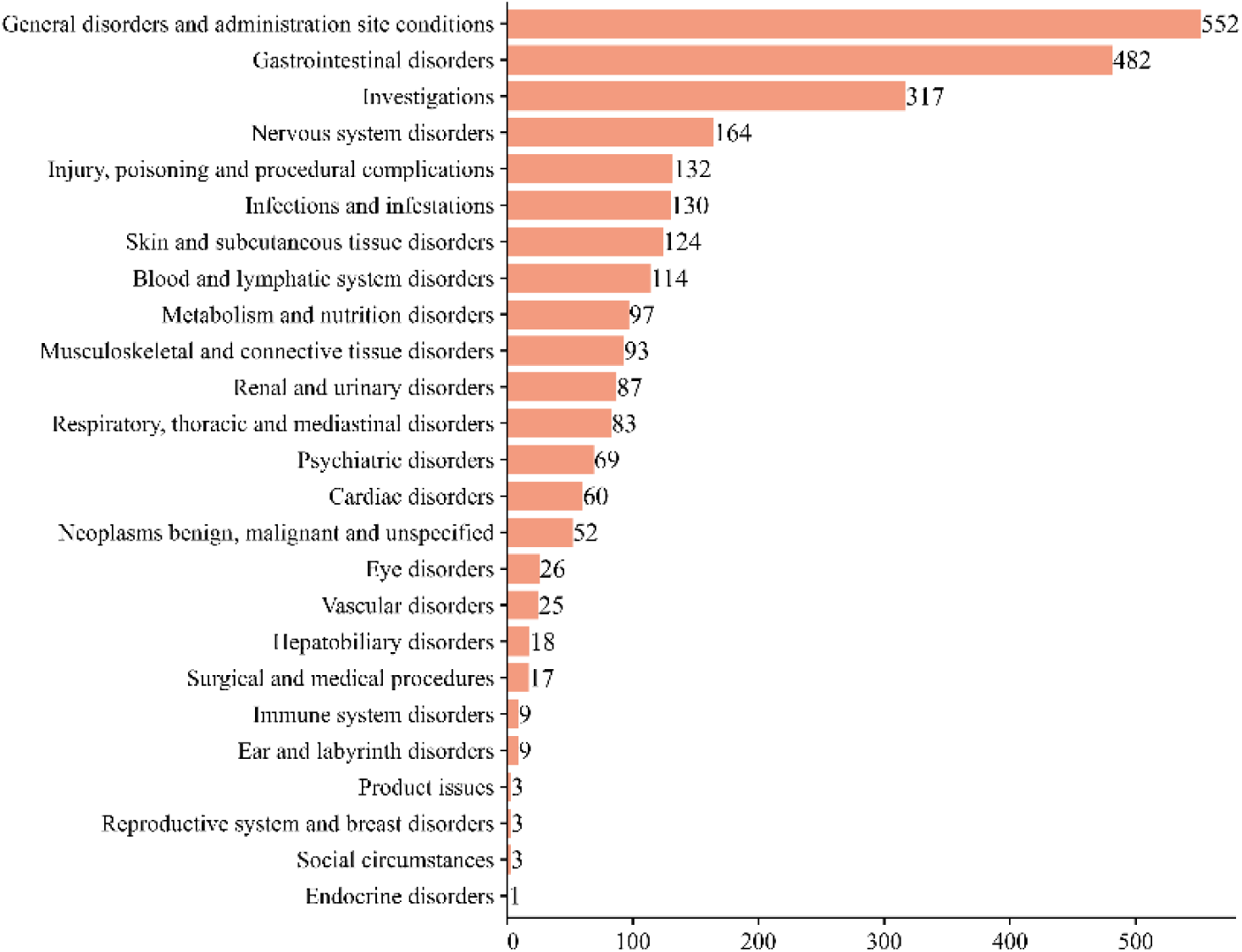
Attribution and number of Fedratinib at the SOC level in the FAERS database.

**Table 3.**
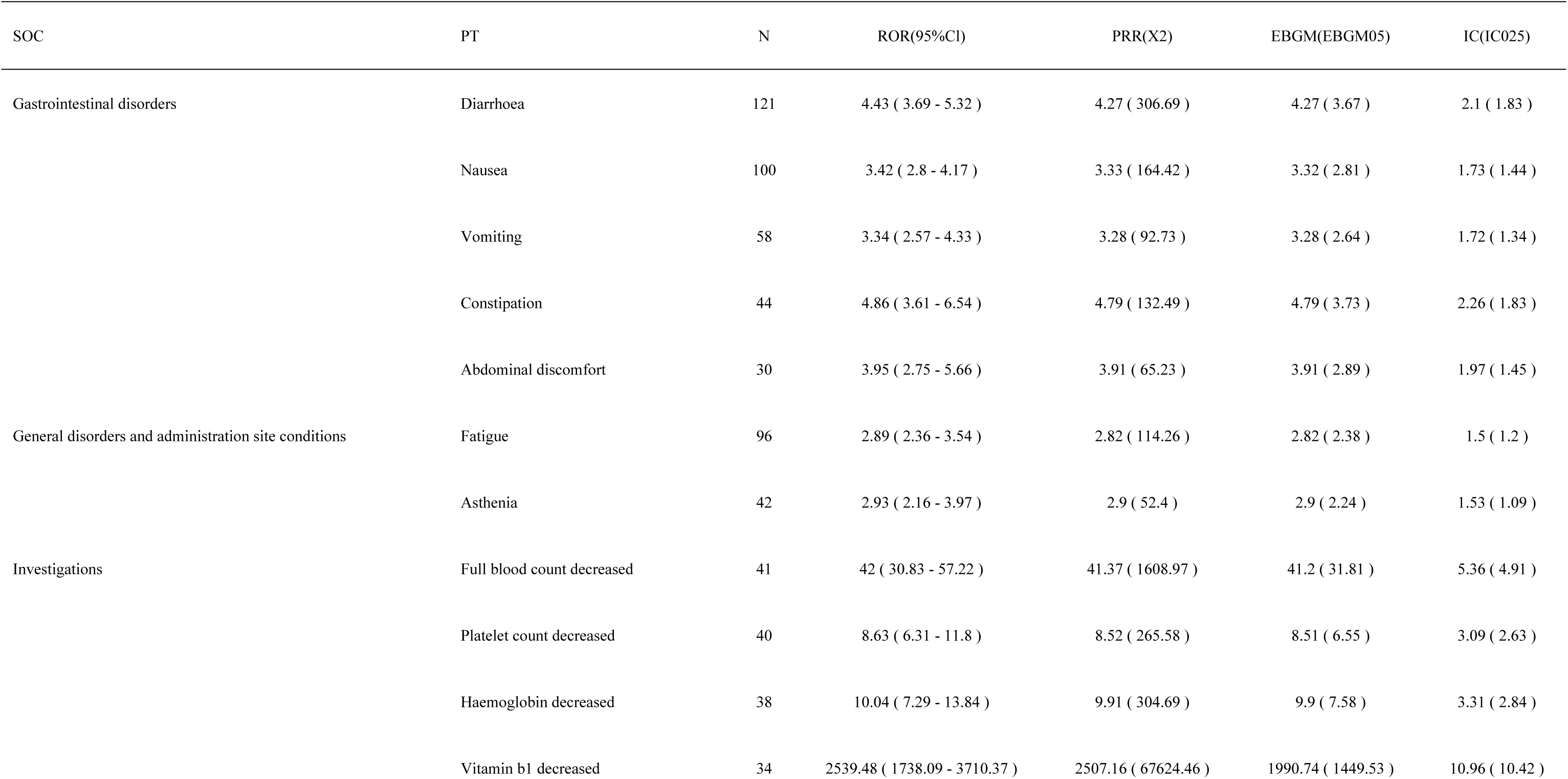
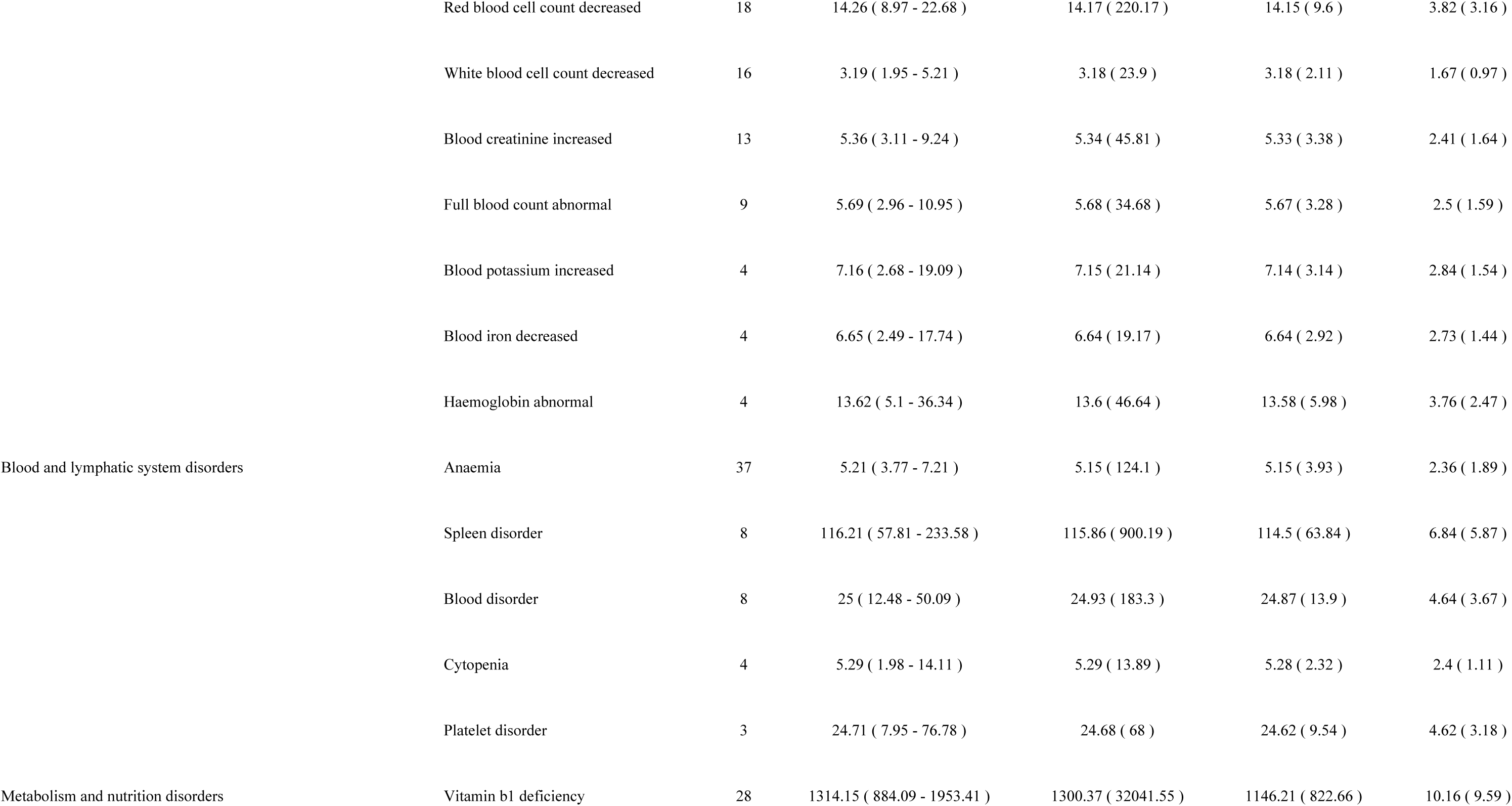
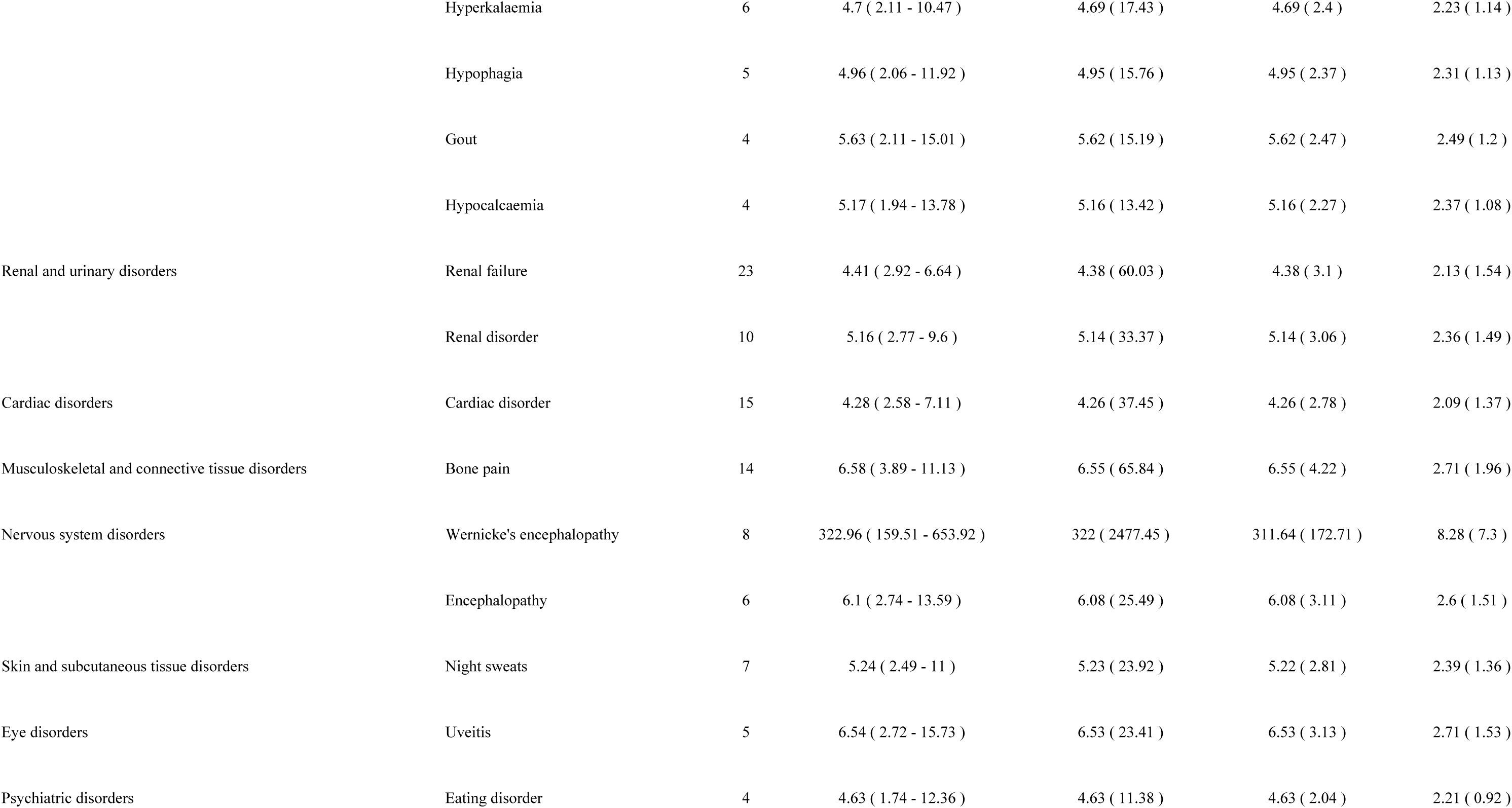
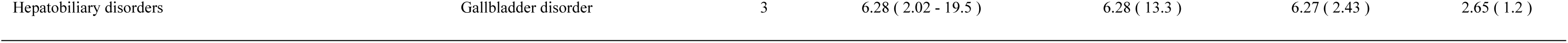
Signal strength of reports of Fedratinib at the PT level in FAERS database.

We ranked the AEs that simultaneously satisfied all four asymmetry analysis methods separately and identified 914 AEs caused by Fedratinib, including 38 PTs that met the screening criteria, corresponding to 13 SOCs (Table 3). The most frequently reported adverse reactions were diarrhoea, nausea, vomiting, fatigue, and constipation. These adverse reactions were consistent with the manufacturer’s prescribing information and established clinical trial findings. Moreover, significant safety signals were detected for several AEs: vitamin b1 decreased (ROR=2539.48, PRR=2507.16, EBGM=1990.74, IC=10.96), vitamin b1 deficiency (ROR=1314.15, PRR=1300.37, EBGM=1146.21, IC=10.16), Wernicke’s encephalopathy (ROR=322.96, PRR=322, EBGM=311.64, IC=8.28), spleen disorder (ROR=116.21, PRR=115.86, EBGM=114.5, IC=6.84), full blood count decreased (ROR=42, PRR=41.37, EBGM=41.2, IC=5.36). In addition, new AEs were identified compared to the most recent package insert issued by the FDA, such as blood potassium increased (ROR=7.16, PRR=7.15, EBGM=7.14, IC=2.84), hyperkalaemia (ROR=4.7, PRR=4.69, EBGM=4.69, IC=2.23), gout (ROR=5.63, PRR=5.62, EBGM=5.62, IC=2.49), hypocalcaemia (ROR=5.17, PRR=5.16, EBGM=5.16, IC=2.37), renal failure (ROR=4.41, PRR=4.38, EBGM=4.38, IC=2.13), renal disorder (ROR=5.16, PRR=5.14, EBGM=5.14, IC=2.36), gallbladder disorder (ROR=6.28, PRR=6.28, EBGM=6.27, IC=2.65). Notably, given the limited case counts for multiple PTs, as presented in Table 3, the IC025 metric was additionally incorporated into our analysis to enhance the robustness of the computational outcomes. Higher IC025 values were found in vitamin b1 decreased (n=34, IC025(10.42)), vitamin b1 deficiency (n=28, IC025(9.59)), Wernicke’s encephalopathy (n=8, IC025(7.3)), spleen disorder (n=8, IC025(5.87)) signals were found to have high IC025 values, although the number of cases under these signals was small, it indicated a close association with Fedratinib administration and deserved our special attention.

In order to highlight the PTs under different SOCs, we use the forest map for visual presentation, as shown in Table 3 and Figure 3. The ROR values and confidence intervals of the different PTs under each system were greater than 1, indicating that these PT signals were significantly and statistically associated with the corresponding SOC. Notably, the corresponding full blood count decreased under investigations showed the strongest correlation, with a significant signal intensity of ROR 42 ( 30.83-57.22 ). We recommend regular assessment of complete blood cell counts as part of routine monitoring for patients undergoing Fedratinib treatment.

**Figure 3.**
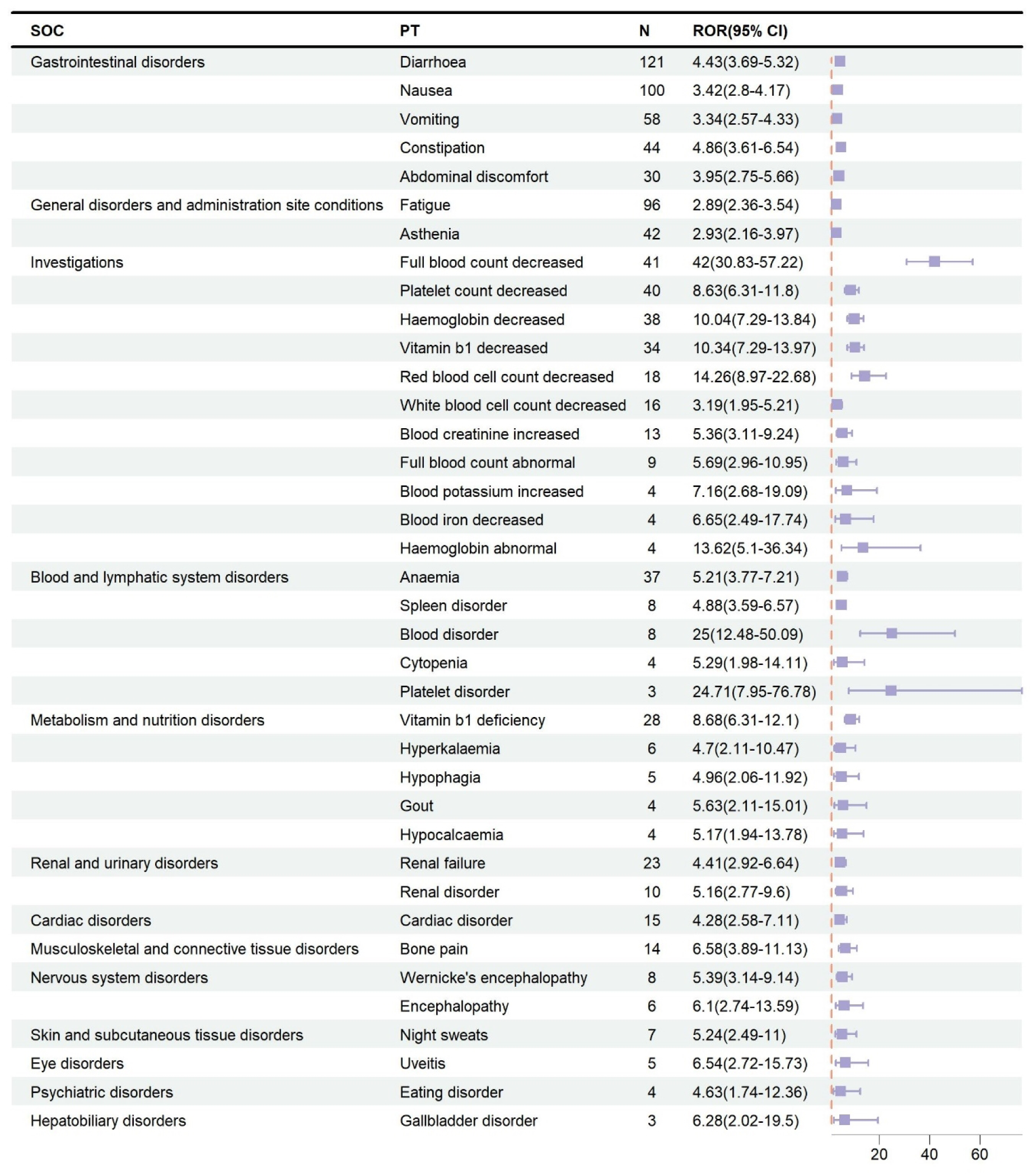
Forest plots for different signal strengths under the ROR algorithm.

### 3.3 Time to onset analysis

The database provided comprehensive documentation of AE onset timelines associated with Fedratinib therapy. Following the exclusion of reports with implausible, incomplete, or unspecified temporal data, 263 quantifiable AE onset intervals were retained for analysis. As illustrated in Figure 4, the majority of cases (n=104, 39.54%) manifested within the initial treatment month, while the number of cases that occurred in the second month decreased to 41 (15.59%), with similar proportions occurring in months 3-6 (4.56%-7.22%), and the incidence of AEs increased during the period of six months to one year after treatment (n=32, 12.17%). It is notable that delayed-onset AEs were observed even after one year of Fedratinib therapy, representing 8.75% (n=23) of the total reported cases. These results underscore the necessity of vigilant adverse event monitoring during Fedratinib therapy. To assess temporal variations in the risk profiles associated with Fedratinib-related AEs, we performed a WSP test on the overall patient population. In this study, we only considered and discussed parameter β. According to the analysis of the results in Table 4, the derived Weibull shape parameter (β) was 0.81 (95% CI upper limit: 0.88), with both values falling below the threshold of 1. This statistical pattern corresponds to the early failure pattern, suggesting a temporally decreasing incidence of Fedratinib-associated AEs.

**Figure 4.**
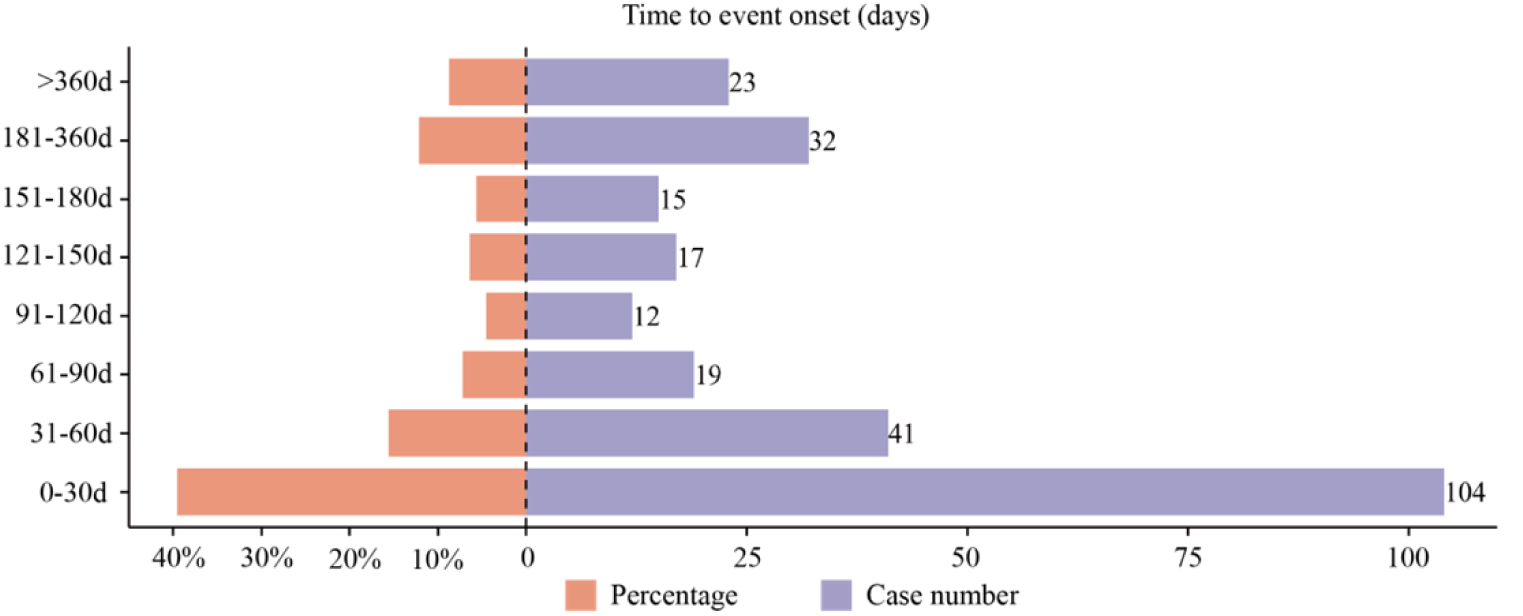
Time to onset of Fedratinib-related AEs.

**Table 4.** Analysis of Weibull distribution of Fedratinib-induced AEs.

|  | coef | 95%CI |
| --- | --- | --- |
| shape parameter: $\beta$ | 0.81 | 0.73-0.88 |
| scale parameter: $\alpha$ | 105.53 | 88.79-122.26 |
| Type | Early failure |  |

### 3.4 Serious vs. non-serious reports

Coincidentally, both SAEs and NSAEs were reported and documented in the current study in 642 cases (50.0% vs. 50.0%). A subgroup analysis of gender and age revealed (Table 5) that male patients demonstrated a higher prevalence of SAEs than female patients (55.0% vs. 45.0%). Male gender showed a non-significant trend toward higher SAE risk compared to females (OR=1.174, 95% CI: 0.939-1.467, p=0.159). In addition, patients aged > 65 years exhibited a significantly higher risk of experiencing SAEs than those aged ≤65 years (≥65 years of age: OR=1.537, 95% CI: 1.102-2.143, P=0.011).

**Table 5.** Comparison of patient gender and age between serious and non-serious reports.

|  | Serious/Non-serious, n | P-value | OR (95%CI) |
| --- | --- | --- | --- |
| Gender |  |  |  |
| Female (Reference) | 274/311 |  |  |
| Male | 334/323 | 0.159 | 1.174 (0.939 - 1.467) |
| Age |  |  |  |
| ≤ 65 (Reference) | 82/97 |  |  |
| > 65 | 365/281 | 0.011 | 1.537 (1.102 - 2.143) |

## 4. Discussion

Previous investigations have predominantly looked at Fedratinib’s pharmacological mechanisms and clinical trial outcomes, and current literature demonstrates a paucity of recent real-world evidence studies examining this association. Therefore, this study employed systematic pharmacovigilance methods to analyze Fedratinib-related adverse drug reactions in real-world settings using FAERS post-marketing surveillance data. This provides AEs with the accuracy and detail to rationalize clinical medication administration, thereby improving the security of clinical drug therapy.

Through systematic pharmacovigilance analysis, we detected 1,284 signals meeting preliminary significance thresholds. The incidence of AEs demonstrated gender and age-specific variations, with male participants showing marginally higher rates than female participants. The elderly population (≥65 years) exhibited the highest AE prevalence, potentially reflecting the increased MF incidence in older males. This is consistent with a study of real-world treatment patterns and clinical outcomes of Fedratinib in patients with MF, in which patients who started using Fedratinib were male with primary MF, with an average age of 67.7 years[23]. These findings underscore the need for enhanced clinical vigilance regarding Fedratinib-associated AEs, particularly in elderly patients, to mitigate the potentially life-threatening complications of Fedratinib. Among the reporting countries/regions, the United States accounted for the majority of reported cases, which may be due to the earliest availability of Fedratinib in the United States and higher prescription volume, or it may reflect regional variations in pharmacovigilance reporting practices or cultural differences in adverse event recognition, warranting further systematic investigation. It is worth noting that although consumers (33.72 %) are the group reporting the most AEs, medical professionals such as medical doctors, pharmacists, health professionals, and other health professionals reported AEs(65.88%), indicating that the data were reliable. The possibility of both serious SAEs and NSAEs caused by Fedratinib was 50%. Although not all AEs can be directly attributed to Fedratinib treatment, these findings necessitate particular attention to Fedratinib-associated AEs, especially SAEs.

Disproportionality analyses determined that the most common SOCs were general disorders and administration site conditions, gastrointestinal disorders, investigations, and nervous system disorders. The most frequently reported adverse reactions comprised diarrhoea, nausea, vomiting, constipation, abdominal discomfort, fatigue, anaemia, platelet count decreased, and Wernicke’s encephalopathy, which are generally consistent with the frequently cited literature and multiple clinical trials of Fedratinib[24–26], and were confirmed in the current study. The initial clinical profile of Fedratinib was established in a Phase I dose-escalation study, with the most frequently reported AEs being gastrointestinal disorders: nausea, diarrhoea, and vomiting in a dose-dependent manner, and hematologic AEs included anaemia, thrombocytopenia, and neutropenia[27]. Given that Fedratinib is well tolerated and shows evidence of clinical activity, it has advanced to clinical development. In a Phase II dose-ranging trial involving patients with either primary or secondary MF, the most common TEAEs at any level were anaemia, diarrhoea, nausea, fatigue, vomiting, and thrombocytopenia. Hematologic toxicities predominated as grade 3-4 AEs, and severe (grade 3-4) TEAEs occurred in 45% of patients, most frequently anaemia, infection, thrombocytopenia, fatigue, and diarrhoea, with anaemia and nausea being the most prevalent[25]. These findings were corroborated in the Phase III JAKARTA trial (NCT01437787) for myelofibrosis, where anaemia, lymphopenia, and thrombocytopenia were the most common high-grade AEs. The most common AEs leading to Fedratinib discontinuation were thrombocytopenia, heart failure, vomiting, and diarrhoea in 19% and 21% of patients who experienced dose reductions and interruptions of Fedratinib 400 mg, respectively, and 14% of patients who experienced permanent discontinuation of treatment[28]. In the Phase II JAKARTA2 trial evaluating Fedratinib in ruxolitinib-pretreated patients with MF, the most frequently reported all-grade AEs included diarrhoea, nausea, anaemia, vomiting, thrombocytopenia, and constipation. Grade 3-4 AEs were predominantly hematologic, with anaemia and thrombocytopenia being the most common[26].

In general, the treatment with Fedratinib is closely related to gastrointestinal events. These AEs mainly occur during early treatment and decrease over time[28]. Although these AEs are common during Fedratinib treatment, they are largely controlled by dose adjustments and rarely require discontinuation of therapy[29]. Therefore, comprehensive patient education and realistic expectation management are essential for promoting medication compliance. Gastrointestinal toxicities are amenable to effective control through prophylactic measures against nausea and vomiting (e.g., administration of 5-HT3 receptor antagonists) and timely therapeutic intervention for diarrhoea upon initial symptom onset. Although food intake does not substantially alter Fedratinib bioavailability, consumption of a lipid-rich diet could provide gastrointestinal tolerability benefits by mitigating nausea and vomiting[30]. The development of anaemia in MF reflects disease progression and portends poorer survival outcomes, which can worsen when Fedratinib is used; therefore, appropriate treatment is necessary[31,32]. Although the use of JAK inhibitors for MF treatment exacerbates anaemia, there is evidence that this is usually temporary and does not reduce survival rates[28,31,33]. Careful patient management, including red blood cell transfusions, treatment of secondary anaemia, JAK inhibitor dosage adjustment, and monitoring, is recommended for patients with MF who develop anaemia, and these therapeutic approaches may ameliorate anaemia, prevent subsequent disease complications, and improve clinical outcomes[34].

In clinical trials involving more than 600 patients treated with Fedratinib, eight reported cases were consistent with Wernicke’s encephalopathy[35], a rare but severe neurological complication of thiamine (vitamin B1) deficiency. As a result, in November 2013, a clinical hold was imposed on the Fedratinib investigational program because of emerging safety concerns. Following the review of supplemental safety data, the FDA removed the clinical hold in August 2017[36]. A black box warning in the Fedratinib prescribing documents alerts clinicians to the risk of treatment-emergent encephalopathy, notably Wernicke’s encephalopathy. Patients should demonstrate adequate thiamine levels prior to initiating Fedratinib therapy, and thiamine levels should be regularly monitored during treatment according to clinical indications. Upon suspicion of encephalopathy, immediate discontinuation of Fedratinib is warranted, and thiamine injections should be initiated until symptom resolution or improvement and normalization of thiamine levels are achieved. The evaluation of PTs revealed a significant signal strength for AEs associated with Fedratinib use. These findings align with established clinical trial evidence and documented safety risks, including decreased vitamin b1 levels, vitamin b1 deficiency, and Wernicke’s encephalopathy. These results underscore the critical need for routine vitamin b1 monitoring and prophylactic supplementation during Fedratinib therapy. All patients should begin thiamine supplementation prior to or concurrently with the initiation of Fedratinib. Patients with low pre-calibration vitamin b1 levels or those whose vitamin b1 levels do not normalize after supplementation cannot use Fedratinib.

Fedratinib treatment demonstrated significant therapeutic efficacy in JAK inhibitor-naïve MF patients, manifesting as both spleen volume reduction and amelioration of MF-related symptom burden. Following the joint guidelines established by the International Working Group for Myeloproliferative Neoplasms Research and Treatment (IWG-MRT) and European LeukemiaNet (ELN)[37], the primary endpoint in myelofibrosis clinical trials assessing JAK inhibitors was defined as the proportion of patients achieving ≥35% reduction in spleen volume from baseline, as measured by MRI or CT scan at the end of cycle 6 (EOC6)[38–43]. Fedratinib induces robust spleen responses, and multiple clinical studies have demonstrated that treatment with Fedratinib leads to both spleen volume response and spleen size response in patients; all evaluable subjects (defined as those with both baseline and EOC6 assessments) demonstrated measurable spleen volume reduction[44–46]. Extended follow-up data demonstrated sustained therapeutic efficacy, with a ≥50% reduction in spleen volume from baseline maintained at 30 months, while Fedratinib treatment exhibited a favorable long-term safety profile without unexpected AEs[47]. This positions Fedratinib alongside ruxolitinib in achieving clinically meaningful reductions in spleen size in patients with myelofibrosis.

In the current study, we also found many hematology-related PT categories, such as full blood count decreased, red blood cell count decreased, blood creatinine increased, etc. These AEs were concentrated in the two SOCs of investigations and blood and lymphatic system disorders. These findings emphasize the importance of regular laboratory testing during Fedratinib therapy. Clinicians should consider the routine evaluation of laboratory parameters to mitigate the side effects associated with these indicators and improve patient prognosis. Given the frequent hematologic complications such as anaemia and thrombocytopenia, controlling treatment-associated cytopenias presents significant clinical challenges during Fedratinib therapy. Similar to ruxolitinib, Fedratinib demonstrates no significant improvement in disease-associated cytopenias. Analysis pooling data from both JAKARTA study phases showed that thrombocytopenia-related treatment discontinuation occurred in only one participant with baseline thrombocytopenia (platelets <100×10^9^/L)[48]. In MF, thrombocytopenia serves as a significant prognostic marker, correlating with disease progression manifested through disease symptom exacerbation, leukemic transformation, and increased mortality[49]. Fedratinib administration requires immediate discontinuation upon detection of thrombocytopenia (platelet count <50×10⁹/L), with therapy reinstatement permissible following platelet recovery to levels exceeding this predefined threshold[50]. Importantly, Fedratinib administration should be avoided when baseline platelet levels fall below 50×10⁹/L due to bleeding risk concerns, which remains a significant unmet clinical need in contemporary hematologic practice[51].

Additional PTs specified in the prescribing information, including hypophagia, cardiac disorders, bone pain, night sweats, and uveitis, etc., were all detected with disproportionality signals in our analysis, further validating the reliability of the dataset. Our pharmacovigilance analysis uncovered multiple previously unreported adverse event signals, such as “blood potassium increased,” “hyperkalaemia,” “gout,”“hypocalcaemia,” “renal failure,” “renal disorder,” and “gallbladder disorder,” none of which were documented in the product labeling or clinical trial reports, and the exact mechanisms responsible for these AEs have not yet been fully defined. The present results emphasize the necessity for continuous monitoring and systematic reporting of AEs, especially those unanticipated in initial clinical trials. Our findings contribute to the growing body of evidence underscoring the necessity of ongoing surveillance and further investigation of this association.

Despite the advantages of data mining techniques in analyzing real-world clinical safety issues, there are some inherent limitations to our study that must be acknowledged. The voluntary and spontaneous reporting system of FAERS has inherent limitations, including potential underreporting, data incompleteness, inaccuracies, inconsistencies, and reporting delays in AEs documentation. Mild AEs and those with a high background prevalence are particularly susceptible to underrepresentation in pharmacovigilance databases, whereas severe or uncommon occurrences could potentially be overestimated in reporting. Second, subjectivity and reporter bias may affect the reporting process, introducing reporting bias. Therefore, these limitations warrant careful consideration in terms of data interpretation and clinical applications. Finally, the identified signals demonstrated statistically significant associations; however, establishing definitive causal relationships requires controlled clinical studies and mechanistic research.

Leveraging the FAERS, we conducted a comprehensive pharmacovigilance analysis of Fedratinib-associated AEs to enhance medication safety in clinical practice, finding substantial consistency between identified AEs and those in the prescribing information while confirming a strong correlation with vitamin b1 deficiency and Wernicke’s encephalopathy. Our systematic assessment characterized Fedratinib’s safety profile through signal detection and temporal analysis, revealing both expected AEs (vitamin b1 decreased, spleen disorder, full blood count decreased) and new signals (blood potassium increased, gout, etc.), underscoring the need for vigilant monitoring across all treated patients. These findings highlight the importance of continuous real-world safety surveillance and warrant further validation through prospective cohort studies and extended clinical trials to precisely quantify Fedratinib-associated risks, as early detection and intervention for both known and unexpected AEs remain crucial for comprehensive pharmacovigilance.

## Statements and Declarations

### Ethics Statement

Ethical approval was not required for this study, as the data analyzed were de-identified records from publicly available database.

### Consent to Participate

Not applicable.

### Consent for Publication

Not applicable.

## Acknowledgments

This study was performed using the FAERS database provided by the FDA. The information, results, or interpretation of the current study do not represent any opinion of the FDA.

## Code Availability

Not applicable.

## Data availability statement

This study analyzed publicly available datasets, which can be accessed here: [https://fis.fda.gov/extensions/FPD-QDE-FAERS/FPD-QDE-FAERS.html].

## Funding

This work was supported by the National Natural Science Foundation of China (No.82460037), the Guangxi Natural Science Foundation (No.2023GXNSFBA026334), the Guangxi Young Elite Scientist Sponsorship Program (No.GXYESS2025185) and the Guangxi High-Level Medical Talent Sponsorship Program.

## Author contributions

LW, YW and YH designed the study. LW, YW, DJ, LT and PZ analyzed the data. JL and QZ made the relative statistics. YW, YL and YX drew the pictures. LW drafted the manuscript and collected the references. YH revised the manuscript. DJ and YH Provided financial support. All authors read and approved the final manuscript.

## Declaration of interests

The authors declare no conflicts of interest or financial ties to any organization related to the manuscript’s subject matter or materials, including employment, consulting, payments, stock ownership, or options.

## Notes

### Competing Interest Statement

The authors have declared that no competing interests exist.

### Funding Statement

The author(s) received no specific funding for this work.

### Author Declarations

Fourth Affiliated Hospital of Guangxi Medical University

